# Outcomes of SARS-CoV-2 Omicron infection in residents of Long-Term Care

**DOI:** 10.1101/2022.01.21.22269605

**Authors:** Maria Krutikov, Oliver Stirrup, Hadjer Nacer-Laidi, Borscha Azmi, Chris Fuller, Gokhan Tut, Tom Palmer, Madhumita Shrotri, Aidan Irwin-Singer, Verity Baynton, The COVID-19 Genomics UK (COG-UK) consortium, Andrew Hayward, Paul Moss, Andrew Copas, Laura Shallcross

## Abstract

**Background:** Recently there has been a rapid, global increase in SARS-CoV-2 infections associated with the Omicron variant (B.1.1.529). Although severity of Omicron cases may be reduced, the scale of infection suggests hospital admissions and deaths may be substantial. Definitive conclusions about disease severity require evidence from populations with the greatest risk of severe outcomes, such as residents of Long-Term Care Facilities (LTCFs).

**Methods:** We used a cohort study to compare the risk of hospital admission or death in LTCF residents in England who had tested positive for SARS-CoV-2 in the period shortly before Omicron emerged (Delta dominant) and the Omicron-dominant period, adjusting for age, sex, vaccine type, and booster vaccination. Variants were confirmed by sequencing or spike-gene status in a subset.

**Results:** Risk of hospital admission was markedly lower in 1241 residents infected in the Omicron-period (4.01% hospitalised, 95% CI: 2.87-5.59) compared to 398 residents infected in the pre-Omicron period (10.8% hospitalised, 95% CI: 8.13-14.29, adjusted Hazard Ratio 0.50, 95% CI: 0.29-0.87, p=0.014); findings were similar in residents with confirmed variant. No residents with previous infection were hospitalised in either period. Mortality was lower in the Omicron versus the pre-Omicron period, (p<0.0001).

**Conclusions:** Risk of severe outcomes in LTCF residents with the SARS-CoV-2 Omicron variant was substantially lower than that seen for previous variants. This suggests the current wave of Omicron infections is unlikely to lead to a major surge in severe disease in LTCF populations with high levels of vaccine coverage and/or natural immunity.

**Trial Registration Number:** ISRCTN 14447421

## Background

The novel B.1.1.529 SARS-CoV-2 variant was first detected in South Africa and was designated a variant of concern named Omicron by WHO on November 26, 2021.^1^ The variant has a large number of mutations in the spike gene, raising concerns about the effectiveness of available vaccines and antibody therapeutics. ^2^ In the 8 weeks since the variant emerged there has been a sharp increase in SARS-CoV-2 infections in all WHO regions, ^3^ and Omicron now accounts for more than 98% of sequenced samples in the UK and USA, ^4,5^ and more than 60% of sequenced samples globally. ^6^ This rapid growth in infections is likely due to the variant’s increased transmissibility ^7^ and its ability to evade immunity conferred by previous infection or vaccination. ^2^

Residents of Long-Term Care Facilities (LTCF) are among the most frail and vulnerable members of society and have been disproportionately affected by the pandemic. An estimated 50% of residents in LTCFs are aged > 85 years,^8^ with high levels of comorbidity. ^9,10^ In the UK, despite high-levels of vaccine coverage in residents (85% have received a booster vaccine),^11^ there has been a rapid increase in the number of outbreaks in LTCFs since December 2021, coinciding with the emergence of the Omicron variant and a rapid increase in SARS-CoV-2 cases nationally.^12^ To date, mortality rates among residents have remained stable, but delays in coding for death certification means that this is a lagged indicator of disease severity. ^13^

Studies in the general population ^14,15^ suggest that the risk of severe outcomes following infection with Omicron may be lower than that seen for previous variants such as Delta, and this risk is attenuated further in those who have received a booster vaccination. ^15–17^ However, the scale of infection suggests that the total number of hospital admissions and deaths due to Omicron may still be substantial, depending on the extent to which age and comorbidity influence disease severity. Data are limited on outcomes following infection in older populations with high rates of comorbidity. In this study, we aimed to investigate the risk of severe outcomes in LTCF residents infected with the SARS-CoV-2 Omicron variant.

## Methods

### Study design

We used a cohort study to investigate the risk of hospital admission and death in residents of LTCFs who tested positive for SARS-CoV-2 between September 1, 2021, and January 17, 2022, and were participating in the VIVALDI study (ISRCTN 14447421). ^18^ We compared outcomes in residents who were infected in the pre-Omicron period when the Delta variant was dominant and, in the Omicron-dominant period. As all LTCF residents in England are screened regularly for SARS-CoV-2, the risk of bias in our assessment of disease severity is relatively low.

Risk of hospital admission was also investigated in a sub-cohort of cases with probable or confirmed Omicron or Delta infections determined by sequencing or S-gene.

### Data Sources

As part of the national testing programme residents in LTCFs in England undertake monthly asymptomatic testing for SARS-CoV-2 using either PCR or Lateral Flow Devices (LFD). They are also tested if they develop symptoms, during outbreaks, or on admission to hospital. ^19^ Each test is linked to a unique identifier based on the individuals’ National Health Service (NHS) number, which can be used to link to other routine datasets.

PCR/LFD test results were linked to national hospital admission and mortality records using the NHS-number based pseudo-identifier. Both of these datasets include International Classification of Diseases 10th edition diagnostic codes. On the date of data extraction (January 17, 2022), admissions data had last been updated on January 14, 2022, and mortality data on January 7, 2022. Vaccine type, and receipt of first or second dose or booster vaccines dose was retrieved by linkage to the National Immunisation Management System (NIMS). LTCF size was retrieved from the Capacity Tracker dataset (https://www.necsu.nhs.uk/capacity-tracker). Data linkage was undertaken securely in the COVID-19 Datastore (https://data.england.nhs.uk/covid-19/).

PCR testing was performed in a network of accredited laboratories established through the national testing programme and a subset of samples were sequenced at the Wellcome Sanger Institute. We retrieved viral lineage for sequenced samples from a publicly available repository, which is established and maintained by the COVID-19 Genomics UK (COG-UK) consortium (https://www.cogconsortium.uk/tools-analysis/public-data-analysis-2/). If sequencing was unavailable, PCR Cycle threshold (Ct) values were used to identify S-Gene Target Failure ‘SGTF’ – a reliable marker of Omicron.^15,20^ Samples with Ct values >30 were excluded from the assessment of SGTF to reduce the risk of misclassifying samples with low viral load, Figure S1. Omicron cases were defined as BA.1 lineage or SGTF. Delta was defined as any AY lineage confirmed on sequencing or detection of S-gene on PCR testing. ^15^

### Study outcomes and covariates

The primary outcome was hospital admission within 14 days following a SARS-CoV-2 positive test, and the secondary outcome was mortality in the 28 days following a positive test. Our main comparison was between two exposure periods based on the date of the first Omicron case in our dataset: the pre-Omicron period when Delta predominated (September 1, 2021 – December 12, 2021) and the Omicron predominant period (December 13, 2021 – January 17, 2022). The comparison of the risk of hospital admission was repeated in the subset of residents with confirmed or probable Delta or Omicron infection based on sequencing or SGTF. Covariates included age (centred at the median), sex, region, LTCF size, prior natural infection (at least one of: previous positive PCR or LFD result in > 28 days before their positive test, prior hospital admission for SARS-CoV-2, or detection of anti-nucleocapsid IgG antibodies). Primary vaccination course was categorised as Pfizer BNT162b, AstraZeneca ChAdOx1, not recorded (in cases where only booster dose was recorded), or unvaccinated. Participants were classified as ‘boosted’ if they had received a third vaccination dose and not boosted if they had received only 2 doses. The time from booster vaccination in days was categorised as follows: no booster before diagnosis, ≤1wk before diagnosis, >1wk before diagnosis).

### Statistical Analysis

We estimated the risk of hospital admission (for any cause) in the 14 days following a positive PCR or LFD test, and plotted Kaplan–Meier curves to compare the cumulative incidence of hospital admission in residents who tested positive during the pre-Omicron (Delta-predominant) and Omicron periods. Residents entered the analysis on the date of their positive test and were censored at the earliest of 14 days following diagnosis for hospitalisations, 28 days following diagnosis for deaths, or the latest date in the dataset (January 17, 2022, for hospitalisation; January 7, 2022, for deaths). We investigated whether the comparison of the risk of hospitalisation between the pre-Omicron and Omicron periods was affected by (i.e., modified by) sex, primary vaccine course, booster status, or age through evaluation of interaction terms in Cox models and stratified Kaplan-Meier plots for the categorical variables. The cumulative incidence of hospital admission was also compared between Delta and Omicron in the known variant cohort using Kaplan-Meier curves.

We modelled risk of hospitalisation in the main cohort using mixed-effects Cox proportional hazards regression with added frailty term to account for LTCF-level clustering. Models were adjusted for age, sex, past infection, primary vaccination type, and time from booster vaccination, with exploration for evidence of an interaction with Omicron period for all adjustment variables. A separate model was constructed in the known variant cohort, using robust standard errors to account for LTCF-level clustering due to the small number of events.

Testing for a difference in the Kaplan-Meier curves between pre-Omicron and Omicron periods, and between Delta and Omicron, was based on the log-rank test. Regression results are presented as adjusted Hazard Ratios with 95% confidence intervals. A p value of < 0.05 was considered statistically significant for effect measures. Formal sample size calculation was not undertaken.

All statistical analyses were conducted in Stata version 16.0. The legal basis for data linkage is provided by the Coronavirus (COVID-19): notice under regulation of the 3(4) of the Health Service (Control of Patient Information) Regulations 2002 (COPI notice). ^21^

Research ethical approval for the study was granted by the South Central Hampshire B NHS REC ref: 20/SC/0238.

## Results

### Cohort description

A total of 697,825 tests were performed in 333 LTCFs, of which 143,227 could not be linked to a pseudo-identifier and 121,655 were performed in residents. Overall, there were 1639 new SARS-CoV-2 diagnoses in 246 LTCFs (Table 1, Figures S1 & S2). The median age of residents with infection was 84.6 years (IQR: 78.0-90.3) and one-third were male. 398 (24.3%) infections were diagnosed in the pre-Omicron period and 1241 (75.7%) were diagnosed in the Omicron-dominant period.

**Table 1:** Baseline characteristics and number of hospital admissions in the 14 days following SARS-CoV-2 diagnosis for care home residents with Omicron versus Delta

|  | All infections | Delta* | Omicron* |
| --- | --- | --- | --- |
| **Total** | 1639<sup>§</sup> | 138 | 407 |
| **Age**<br>(IQR, range) | 84.6 (78.0-90.3, 53.0-104.9) | 84.9 (77.9-91.4, 66.0-100.7) | 85.2 (79.0-90.6, 60.0-103.7) |
| **Sex** |  |  |  |
| Female | 1129 (68.9) | 92 (66.7) | 293 (72.0) |
| Male | 510 (31.1) | 46 (33.3) | 114 (28.0) |
| **Time period of positive test** |  |  |  |
| Pre-Omicron: September 1, 2021 - December 12, 2021 | 398 (24.3) | 96 (69.6) | 0 (0.0) |
| Omicron: December 13, 2021 - January 17, 2022 | 1241 (75.7) | 42 (30.4) | 407 (100.0) |
| ***Type of test*** |  |  |  |
| Identified by LFD | 147 (9.0) | 0 (0.0) | 0 (0.0) |
| Identified by PCR | 1492 (91.0) | 138 (100.0) | 407 (100.0) |
| ***Primary vaccine course*** |  |  |  |
| AstraZeneca ChAdOx1 | 851 (51.9) | 69 (50.0) | 216 (53.1) |
| Pfizer BNT162b2 | 487 (29.7) | 39 (28.3) | 130 (31.9) |
| Not recorded | 69 (4.2) | 7 (5.1) | 12 (2.9) |
| Unvaccinated | 232 (14.2) | 23 (16.7) | 49 (12.0) |
| ***Booster vaccination status<sup>‡</sup>*** |  |  |  |
| No booster before positive test | 619 (37.8) | 89 (64.5) | 116 (28.5) |
| Booster ≤ 1 week before positive test | 42 (2.6) | 10 (7.2) | 5 (1.2) |
| Booster > 1 week before positive test | 978 (59.7) | 39 (28.3) | 286 (70.3) |
| ***Days from booster to positive test***<br>(IQR, range) | 75 (55-89, 1-182) | 42 (10-72, 1-89) | 78 (66-92, 1-112) |
| ***Infection history*** |  |  |  |
| Any evidence of past infection | 172 (10.5) | 5 (3.6) | 46 (11.3) |
| Previous positive PCR or LFD | 117 (7.1) | 3 (2.2) | 32 (7.9) |
| SARS-CoV-2 antibodies | 17 (1.0) | 0 (0.0) | 0 (0.0) |
| Prior COVID-19 hospital admission | 71 (4.3) | 2 (1.4) | 22 (5.4) |
| ***Follow-up time hospitalisation (IQR, range)*** |  |  |  |
| Positive test September 1, 2021 -<br>December 12, 2021 | 14 (14-14, 14-14) | 14 (14-14, 14-14) | - |
| Positive test December 13, 2021 -<br>January 17, 2022 | 14 (8-14, 2-14) | 14 (14-14, 7-14) | 12 (6-14, 3-14) |
| ***Follow up time death (IQR, range)*** |  |  |  |
| Positive test September 1, 2021 -<br>December 12, 2021 | 28 (28-28, 26-28) | 28 (28-28, 28-28) | - |
| Positive test December 13, 2021 -<br>January 17, 2022 | 4 (0-10, 0-25) | 18 (16-24, 0-25) | 2 (0-5, 0-25) |
| ***Hospital Admission with 14 days of positive test*** |  |  |  |
| Any | 79 (4.8) | 12 (8.7) | 7 (1.7) |
| Positive test September 1, 2021 -<br>December 12, 2021 | 43 (10.8) | 5 (5.2) | 0 (0) |
| Positive test December 13, 2021 -<br>January 17, 2022 | 36 (2.9) | 7 (16.7) | 7 (1.7) |
| ***COVID-19 deaths<sup>#</sup>*** |  |  |  |
| All | 55 (3.4) | 14 (10.1) | 2 (0.5) |
| Positive test September 1, 2021 -<br>December 12, 2021 | 47 (11.8) | 12 (12.5) | 0 (.) |
| Positive test December 13, 2021 -<br>January 17, 2022 | 8 (0.6) | 2 (4.8) | 2 (0.5) |
| ***Region*** |  |  |  |
| East Midlands | 195 (11.9) | 14 (10.1) | 71 (17.4) |
| East of England | 107 (6.5) | 5 (3.6) | 31 (7.6) |
| London | 172 (10.5) | 0 (0.0) | 32 (8.6) |
| North East | 100 (6.1) | 2 (1.4) | 42 (10.3) |
| North West | 436 (26.6) | 41 (29.7) | 149 (36.6) |
| South East | 133 (8.1) | 24 (17.4) | 11 (2.7) |
| South West | 280 (17.1) | 20 (14.5) | 25 (6.1) |
| West Midlands | 130 (7.9) | 12 (8.7) | 16 (3.9) |
| Yorkshire and The Humber | 86 (5.2) | 20 (14.5) | 27 (6.6) |
| **Number of care home beds** | 58 | 59 | 60 |
| (IQR, range) | (44-80, 17-149) | (45-77, 24-127) | (45-81, 23-149) |

*§5 residents had 2 infections > 28 days apart in the dataset and in both cases the first positive test was dropped from the analysis*
*\*Subset of probable and confirmed cases based on lineage derived from sequencing and/or SGTF for cases of Omicron.*
*#COVID-19 deaths defined as death within 28 days of PCR or COVID-19 recorded on death certificate. Data are incomplete for the Omicron period due to time-lag in data collection.*
*±Booster vaccinations shown for entire study cohort, regardless of primary vaccination status*

In total, 79 residents were admitted to hospital in the 14 days following PCR/LFD-positive infection. This included 43 admissions in 398 residents who were infected in the pre-Omicron period and 36 admissions in 1241 residents who were infected in the Omicron-dominant period. 172 out of 1639 (10.5%) of participants had evidence of past infection however none of these residents were admitted to hospital. There were 48 deaths in the 28 days following infection: 40 occurred in 398 residents infected in the pre-Omicron period compared to 8 deaths in 868 residents infected in the Omicron-predominant period, Table 1.

### Risk of hospitalisation

The incidence of hospital admission following infection was lower for individuals who were infected in the Omicron period (4.01% admitted within 14 days of a positive test, 95% CI: 2.87-5.59) versus the pre-Omicron period (10.80% admitted within 14 days following a positive test, 95% CI: 8.13-14.29), Figure 1, log-rank test p<0.0001. This effect was seen in both men and women (Figures S3A and S3B).

**Figure 1.**
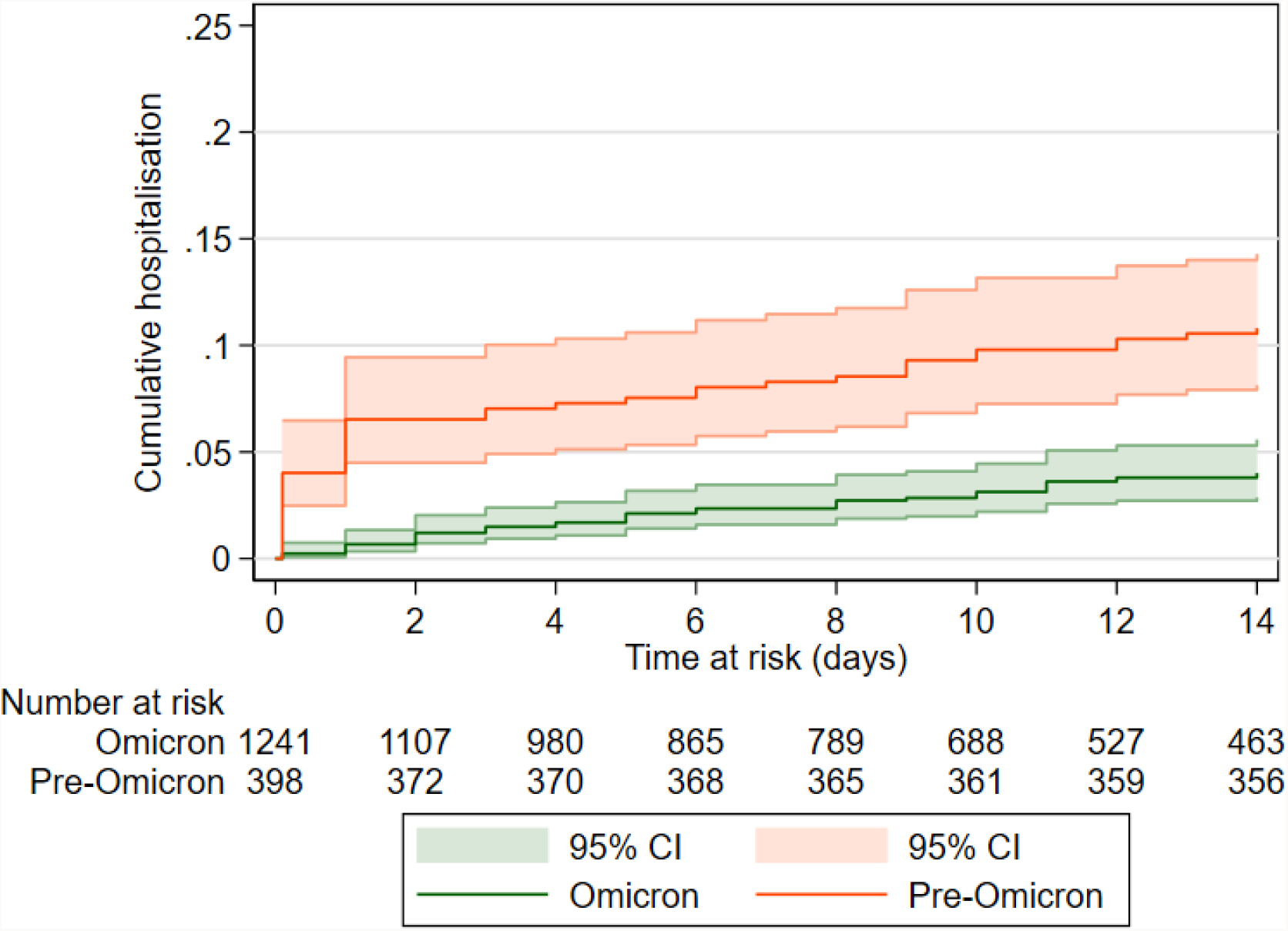
Kaplan Meier curve comparing the cumulative incidence of hospital admission in the 14 days following a positive PCR or LFD test in the pre-Omicron (September 1, 2021 - December 12, 2021) and Omicron periods (December 13, 2021 - January 14, 2022), p<0.0001. Participants who are not hospitalised are censored at the earliest of 14 days following test date or January 14, 2022.

The same pattern of reduced risk of hospital admission in the Omicron versus pre-Omicron period was seen in residents irrespective of whether they had received a primary course of AstraZeneca ChAdOx1 or Pfizer BNT162b2 vaccine, or remained unvaccinated, Figures S4A, S4B, and S4C.

Reduced risk of hospitalisation in the Omicron period was also seen in residents who had received a booster vaccine before their diagnosis, Figure S5A, log-rank test p<0.0001. However, the risk appeared similar in the pre-Omicron and Delta periods in those who had not received a booster, Figure S5B, p=0.549.

The unadjusted Hazard Ratio (HR) for hospitalisation following diagnosis in the Omicron period compared to the pre-Omicron period was 0.35 (95% CI: 0.22-0.55, p<0.0001), and this effect was slightly attenuated in the multivariable model (aHR 0.50 (95% CI: 0.29-0.87, p=0.014). The adjusted risk of hospital admission was lower in women compared to men (aHR 0.61, 95%CI: 0.39-0.97, p=0.037) whereas age and vaccine type were not independently associated with outcome, Table 2. There was evidence of interaction between the risk of hospital admission in the pre-Omicron/Omicron periods and booster vaccine status (p=0.045), with the greatest relative reduction in risk from Omicron in residents who had received booster vaccination > 1 week before diagnosis (aHR 0.23, 95% CI: 0.10-0.53, p=0.001). Interaction was also seen with primary vaccine type (p=0.045), full models shown in Table S1.

**Table 2:** Cox proportional Hazards model for hospitalisation within 14 days from positive test for SARS-CoV-2 in the full cohort and the known variant cohort. Models are adjusted for median-centred age and all other variables listed in the model.

| Variable | Adjusted<br>HR | 95% CI | P value | Adjusted<br>HR | 95% CI | P value |
| --- | --- | --- | --- | --- | --- | --- |
|  | **Overall (n=1467, clusters=240)<sup>#</sup>** |  |  | **Known variant cohort (n=494)<sup>\*</sup>** |  |  |
| **Period / Variant** |  |  |  |  |  |  |
| Pre-Omicron / Delta | Ref | - | - | Ref | - | - |
| Omicron | 0.50 | 0.29-0.87 | 0.014 | 0.33 | 0.11-0.98 | 0.045 |
| **Sex** |  |  |  |  |  |  |
| Male | Ref | - | - | Ref | - | - |
| Female | 0.61 | 0.39-0.97 | 0.037 | 1.68 | 0.59-4.83 | 0.333 |
| **Age (per year increase)** | 1.02 | 1.00-1.05 | 0.111 | 1.02 | 0.96-1.08 | 0.506 |
| **Vaccine type** |  |  | 0.6948 |  |  | <0.0001 |
| Unvaccinated | Ref | - | - | Ref | - | - |
| AstraZeneca ChAdOx1 | 1.06 | 0.51-2.18 | - | 0.45 | 0.09-2.17 | - |
| Pfizer BNT162b2 | 0.89 | 0.45-1.73 | - | 0.24 | 0.06-1.06 | - |
| Type not known | 0.47 | 0.10-2.18 | - | - | - | - |
| **Booster vaccine status<sup>§</sup>** |  |  | 0.0239 |  |  | 0.0758 |
| No booster | Ref | - | - | Ref | - | - |
| Booster within week before Dx | 2.00 | 0.85-4.65 | - | 6.23 | 1.21-32.07 | - |
| Booster more than 1 week before Dx | 0.56 | 0.30-1.06 | - | 1.36 | 0.36-5.16 | - |

*Cases with known past infection have been omitted from model fitting, because of a lack of hospitalisation events in this group.*
*#Frailty term added to adjust for LTCF-level clustering*
*\*Robust standard errors to adjust for LTCF-level clustering*
*§Unvaccinated residents included in model as 'unboosted' but due to inclusion of vaccine type in the model the aHRs for boosting are the additional effects of boosting in those with primary vaccination*

Next, we confirmed our findings in the subset of 407 probable/confirmed Omicron and 138 probable/confirmed Delta infections in 545 residents based on lineage (52 Omicron, 6 Delta) and / or the presence of SGTF (377 Omicron, 134 Delta), Figure S1. In this cohort there were 12 admissions and 2 deaths in 138 residents with Delta versus 7 admissions and 2 deaths in 407 residents with Omicron. The proportion of cases admitted was higher in cases infected with Delta (8.82%, 95% CI: 5.11-15.02) compared to Omicron (3.77%, 95% CI: 1.57-8.95) Figure 2, log-rank test p=0.009. The unadjusted (HR 0.32, 95% CI: 0.11-0.92, p=0.034) and adjusted (aHR 0.33, 95% CI: 0.11-0.98, p=0.045) risks of hospital admission for Omicron versus Delta infections were similar to that seen in the main analysis.

**Figure 2.**
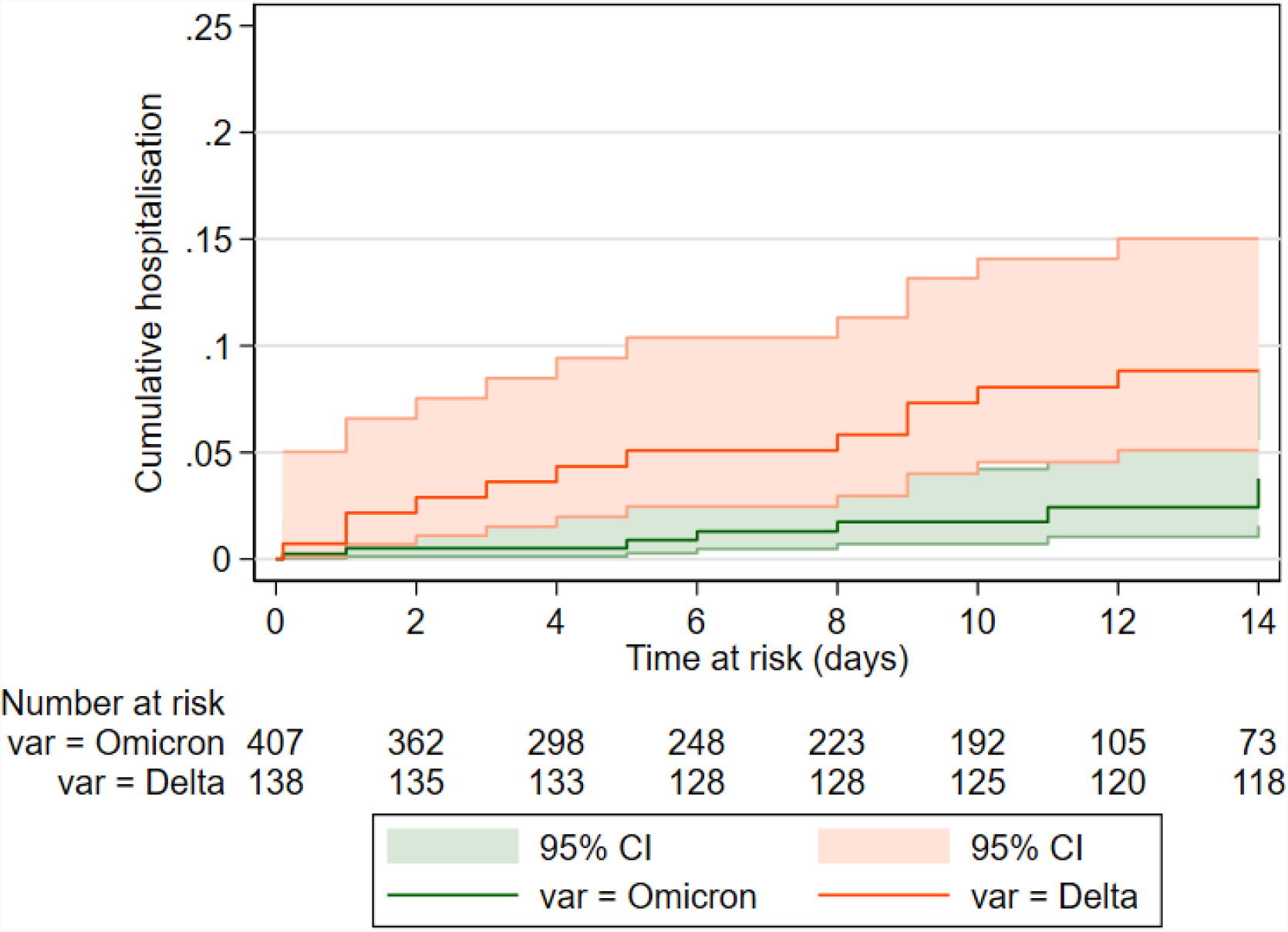
Cumulative incidence (risk) of hospital admission in residents with confirmed/probable Delta infection versus those with confirmed/probable Omicron infection, (p=0.009) based on sequencing and S-gene target failure. Participants who are not hospitalised are censored at the earliest of 14 days following the test date or January 14, 2022.

### Risk of death

There was a reduced risk of death within 28 days of a new SARS-CoV-2 diagnosis in the Omicron-dominant period (1.1 deaths / 1000 person-days, 95% CI: 0.6-2.2,) compared to the pre-Omicron period (3.8 deaths / 1000 person-days, 95% CI: 2.8-5.2), log-rank test p<0.0001, Figure 3. However, further modelling was not performed due to the limited follow-up time in the Omicron-dominant group at this time.

**Figure 3.**
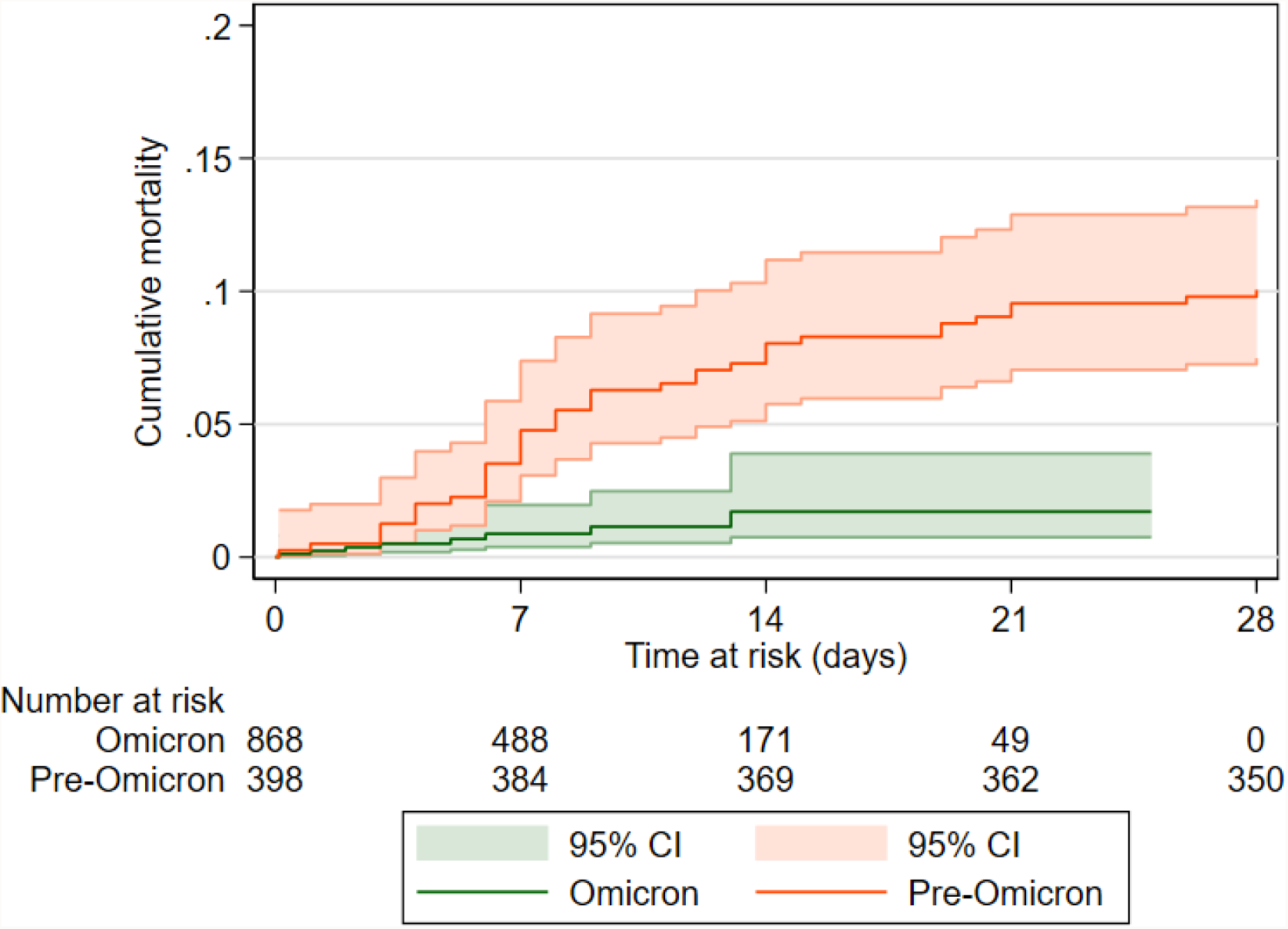
Cumulative mortality in 28 days following SARS-CoV-2 test in the pre-Omicron and Omicron period (p<0.0001), between September 1, 2021 and January 7, 2022. Participants who do not reach the outcome are censored at the earliest of 28 days following the test date or January 7, 2022.

## Discussion

In this study in residents of LTCFs with SARS-CoV-2 infection, we found that disease severity was substantially reduced following the emergence of the Omicron variant. This effect was seen for hospital admissions and mortality, although further follow-up time is required to confirm our findings on mortality. Similar results were obtained when we restricted our analysis to confirmed Delta or Omicron infections, increasing confidence in our findings.

The majority of residents in our study were fully vaccinated, and 62% had received booster vaccination (3rd dose) before they tested positive for SARS-CoV-2. In common with studies in the general population, we found some evidence that receipt of a booster vaccine augmented the reduction in risk of hospitalisation that was seen in the Omicron versus pre-Omicron period. However, it is difficult to disentangle the direct effect of the variant on severe outcomes because our analysis does not account for the overall effect of vaccination on infection, and the impact of waning immunity and the community incidence of infection are likely to have varied in the pre-Omicron and Omicron-dominant periods. ^22–25^

To date, no publications have described severity of Omicron infection in LTCF residents, however preliminary findings from community-dwelling older adults are consistent with the results that we have reported. One large study awaiting peer-review that included >50,000 positive tests in South California in adults older than 65 years reported a lower risk of hospitalisation following symptomatic SGTF (Omicron) compared with non-SGTF (Delta) infections (aHR 0.36, 95% CI: 0.19-0.70). ^26^ Similarly, a matched cohort study in adults aged >60 years in Canada, which differentiated variants using a combination of sequencing, S-gene and onset date, reported a 60% reduction in the risk of hospitalisation or death following Omicron infections compared to Delta infection (aHR 0.40, 95% CI: 0.28-0.56). ^27^

Regular asymptomatic testing for SARS-CoV-2 in residents of LTCFs in England allowed us to obtain a relatively unbiased estimate of disease severity, in contrast to the majority of studies which focus on symptomatic cases. Through the VIVALDI study we were able to link to routine datasets recorded in near-real time which made it possible to reliably capture outcomes in participants and to rapidly assess the impact of Omicron. We also had access to viral lineages obtained through the UK’s large-scale whole genome sequencing programme, which made it possible to confirm variant type in one-third of infections. Our study provides important insights into the risk of severe outcomes in LTCF residents, who are frequently excluded from research studies, and have experienced among the highest rates of SARS-CoV-2 related morbidity and mortality.

Our study has several limitations. First, our ability to assess the effect of Omicron on mortality is constrained by the duration of follow-up in our study to date. Second, as not all laboratories use assays that include the S-gene target, it was not possible to identify all samples with SGTF or to confirm that samples with SGTF were indeed Omicron. However, all 26 samples that had been sequenced and tested for S-gene were concordant. The recent emergence of a sub-variant of Omicron of the BA.2 lineage that does not exhibit SGTF are unlikely to have affected our results as, based on our analysis of data from the UK’s genomic surveillance programme, only 24 of samples of this lineage have been identified to date. Finally, it is likely that we underestimated the prevalence of past infection in our cohort (10.5%), which is significantly lower than published seroprevalence estimates from the LTCF population, ^28^ because only a subset of residents had been tested for antibodies to nucleocapsid.

## Conclusions

Overall, the markedly decreased severity combined with high vaccination uptake and prior natural infection can be expected to significantly limit the impact of the current wave of Omicron infections on hospitalisations and deaths in residents of LTCFs.

## Data Availability

De-identified test results and limited meta-data will be made available for use by
researchers in future studies, subject to appropriate research ethical approvals, once the
VIVALDI study cohort has been finalised

## Contributors

LS, AC, OS, and MK conceptualised the study. MK, OS, LS, and AC developed the statistical analysis plan. MK and OS did the formal statistical analysis. MK, CF, BA, HNL, RB, MS and AI-S were involved with project administration. LS and AH obtained research funding. MK and LS wrote the first draft of the manuscript. All authors revised and edited the manuscript. MK and OS accessed and verified the data. All authors had full access to all the data reported in the study. LS, AC, and MK shared the final responsibility for the decision to submit for publication.

## Data sharing

De-identified test results and limited metadata will be made available for use by researchers in future studies, subject to appropriate research ethical approvals once the VIVALDI study cohort has been finalised. These datasets will be accessible via the Health Data Research UK Gateway.

## Acknowledgements

We thank the staff and residents in the LTCFs that participated in this study and Mark Marshall at NHS England who pseudonymised the electronic health records.

